# COVID-19 Vaccination and Healthcare Demand

**DOI:** 10.1101/2021.09.15.21263628

**Authors:** Matthew I. Betti, Amira Hassan Abouleish, Victoria Spofford, Cory Peddigrew, Alan Diener, Jane M. Heffernan

## Abstract

One of the driving concerns during any epidemic is the strain on the healthcare system. As we have seen many times over the globe with the COVID-19 pandemic, hospitals and ICUs can quickly become overwhelmed by cases. While strict periods of public health mitigation have certainly helped decrease incidence and thus healthcare demand, vaccination is the only clear long-term solution. In this paper, we develop a two-module model to forecast the effects of relaxation of non-pharmaceutical intervention and vaccine uptake on daily incidence, and the cascade effects on healthcare demand. The first module is a simple epidemiological model which incorporates non-pharmaceutical intervention, the relaxation of such measures and vaccination campaigns to predict caseloads into the the Fall of 2021. This module is then fed into a healthcare module which can forecast the number of doctor visits, the number of occupied hospital beds, number of occupied ICU beds and any excess demand of these. From this module we can also estimate the length of stay of individuals in ICU. For model verification and forecasting, we use the four most populous Canadian provinces as a case study.

## 1 Introduction

COVID-19 continues to strain healthcare systems globally [38, 24, 35, 3]. To combat the spread of SARS-CoV-2, many countries have implemented various public health mitigation measures (i.e., quarantine, social distancing recommendations, schools closures, border closures) and have cancelled elective healthcare procedures to reduce healthcare demand and occupancy [22, 27, 8]. Vaccination campaigns have also been launched in ways to minimize healthcare demand, vaccinating healthcare workers and those with the highest potential for severe outcome first [31, 1, 20].

In Canada, periods of strict public health mitigation have certainly worked to decrease infection, and thus, healthcare and hospital demand (see Figure 1, left panel). COVID-19 vaccination in Canada started out slow, but it accelerated through Spring and early Summer 2021 (see Figure 1, right panel).

**Figure 1:**
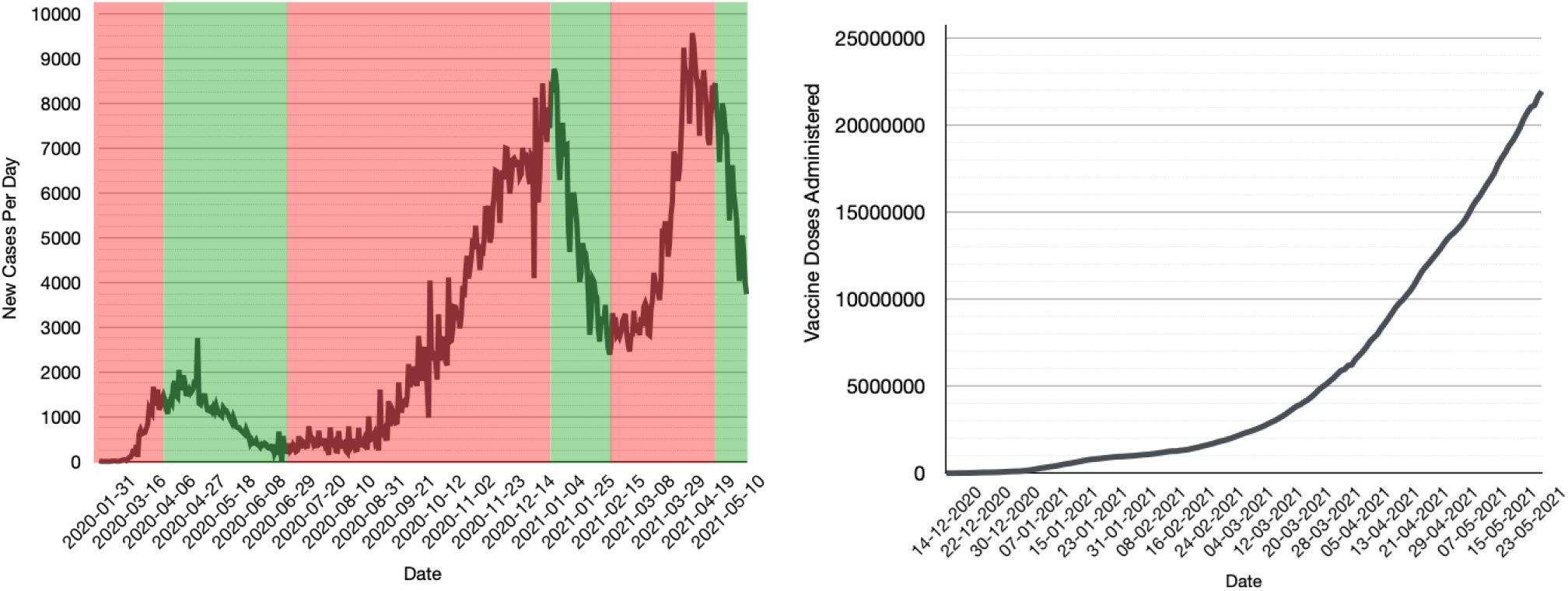
COVID-19 cases and vaccination in Canada. (left) New reported cases per day. The green shaded regions generally correspond to strict public health lockdowns in Canada’s three largest provinces (Ontario, Quebec, British Columbia). The red shaded region is the time period after a relaxation of public health measures (*e*.*g*. reopening of schools, non-essential retail, construction, *etc*.). (right) Total vaccine doses administered. We see that by May 23, 2021 roughly 57% of Canadians have had at least one dose of a COVID-19 vaccine. Note the slow start of the vaccination program in late 2020 into early 2021.

Until June 2021, the Canadian COVID-19 vaccination program focused on accelerated delivery of one dose of vaccine to as many Canadians as possible [16, 36], so as to minimize healthcare demand and allow for some relaxation of public health mitigation programs for Summer 2021. Delivery of the second dose of vaccine in two-dose vaccine regimens was to be a focus over Summer 2021 and into the Fall, but earlier delivery of the second dose was initiated in response to the *δ*-variant. It has been observed that vaccination rates experience decreases in uptake once a country has reached > 40 − 50% coverage (see, for example, the vaccine uptake for Canada, Hungary, Israel, United Kingdom, United States of America at [37]). It has also been observed that lower infection rates can induce relaxation in personal protective behaviours (i.e., social distancing and mask wear). Canada achieved 50% population coverage of at least once dose in mid-May 2021. A reduction in vaccine uptake over the summer months unfortunately occurred [37]. Relaxation in personal protective behaviours also occurred over Summer 2021, particularly when provincial jurisdictions relaxed restrictions and opened borders (in early July 2021). Reductions in vaccination uptake and protective behaviour will affect the probability and degree of Fall 2021 COVID-19 resurgence. The expected burden on the healthcare system needs to be quantified.

Mathematical models have been used to look at various aspects of the COVID-19 epidemic in Canada and globally. Early models focused on parameter estimation and effects of different mitigation strategies on disease spread [26, 39]. More recently, models have looked at the correlation between mobility patterns and disease transmission [2], or the interplay between vaccination and relaxing the aforementioned mitigation strategies [32, 6]. While there have been studies on the impact of COVID-19 on healthcare infrastructure in various countries [15, 30, 14], there has been no such study for Canada as of yet. Moreover, there is a need for a generalized framework from which to build such models that can be adapted to different regions.

We have developed a mathematical framework to study healthcare demand in Canada, adapted from a model developed by [34, 33] for Australia. The framework consists of two modules: a mathematical epidemiology module that consists of mathematical model of COVID-19 infections (mild and severe, reported and unknown); and a healthcare demand clinical pathways module adapted from [34, 33], through which outcomes of the epidemiological model are fed, in order to quantify healthcare demand. Here within, we employ our mathematical framework to study scenarios of COVID-19 infection in Summer and Fall 2021, with lower vaccination uptake and social distancing behaviour.

## 2 Model

The modelling framework presented here consists of two modules, a mathematical modelling module and a clinical pathways module, that include models of COVID-19 epidemiology and healthcare demand. The modules are used in combination to project COVID-19 infection load during a specified time period, and determine the need for healthcare resources i.e., doctor (GP) and emergency room (ED) visits, COVID-19 assessment clinic visits, and ward (non-ICU) and ICU beds.

### 2.1 Mathematical Epidemiology Module

The clinical pathways module is informed by an underlying epidemiological model. The clinical pathway model requires cases to be separated into mild and severe cases; these are defined as cases with no/mild/moderate symptoms, and severe symptoms with hospitalization needed, respectively. In a previous study [7], we developed an epidemiological model that (1) is simple enough, with few enough parameters that we can fit the model to small data sets, and (2) separates mild and severe cases, that enables model results to be fed into the clinical pathways model. Herewithin, we employ this model in the Mathematical Epidemiology Module.

Briefly, our model tracks four compartments: mild active cases, *I*_*m*_, severe active cases, *I*_*s*_, cumulative known cases, *C*_*K*_, and cumulative total cases (i.e. total incidence), *C*_*I*_. The governing equations are

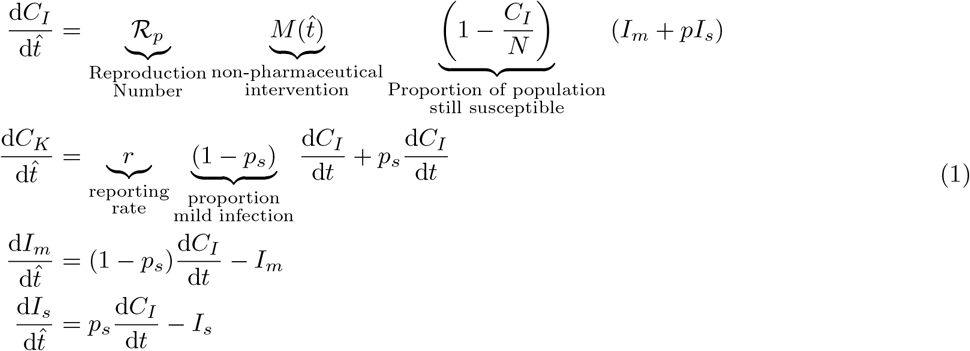

Here, ℛ_*P*_ is the population reproduction number, *p* is a scaling factor on severe cases which limits their ability to spread disease (increased non-pharmaceutical interventions placed on severe cases), *p*_*s*_ is the probability of an infection being severe, *r* is the reporting rate, and *M* (*t*) is a mitigation function which can be tuned to account for increases and relaxations of non-pharmaceutical interventions (NPI). For the model, time is measured in infectious lifetimes, 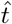.

The mitigation function takes on the general form

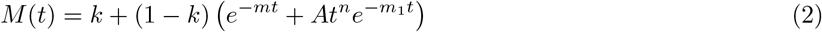

which allows for both non-pharmaceutical interventions and relaxation of said interventions.

The model is subject to a number of assumptions:

1. The total population is constant.
2. Acquired immunity lasts longer than the outbreak.
3. There is no co-infection or super-infection.
4. The testing/reporting rate in the population is relatively constant.
5. The probability of a case being severe vs. mild is constant.
6. All severe infections are reported, whereas a fraction of mild infections are.

We see from these assumptions that the model is best used for short-to moderate-term forecasting (particularly assumptions 1 and 2).

We fit Model (1), with Equation (2), using a least squares method on the cumulative known cases. In the current study, the model is fit from February 15, 2021 to May 18, 2021, the date corresponding to 50% population coverage of at least one dose of vaccine. In order to maximize the number of data points used, we use both cumulative known cases and new known cases per day (i.e. the derivative of cumulative known cases). We minimize the error function

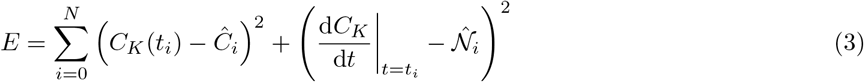

where *Ĉ*_*i*_ is the cumulative known case measurement on day *t*_*i*_ and 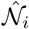 is the new cases measured on day *t*_*i*_. We use a modified bootstrapping method which is described in [7]. We run the least squares fitting on different random subsets of the data, and use the mean of all fits as the fit. This process is similar to how a random forest generates classifications and reduces overfitting [28].

The model fit for each province is shown in Figures 3 to 6, depicted by the blue line. The model is carried forward past the last day of data considered in the model fitting (May 18, 2021, orange vertical line). It is in close agreement with case data for each province past this date to June 1. In all provinces the blue line reaches a daily incidence of approximately zero by early August 2021. However, NPI relaxation, changes to vaccine efficacy due to introductions of new variants of concern, and lower uptake of the vaccines, will affect this outcome.

We note that Model (1) does not explicitly model vaccination. The case data, however, incorporates effects of vaccine uptake. In the model fit, the effects of vaccination are reflected in the fitted value of parameter ℛ_*P*_ which incorporates effects of vaccination i.e., reductions in the average susceptibility of susceptibles (including vaccinated and non-vaccinated individuals), and reductions in transmission capabilities of infecteds that were vaccinated.

Once we have the baseline fit (from February 15, 2021 to May 18, 2021), we are able to explore different scenarios which incorporate dynamics that explicitly consider increases in protection from infection in vaccinateds, and in the relaxation of NPI measures. Beginning June 1 2021, we would like to consider modifications to the effects of vaccination and NPI relaxation in the system. In (4) we modify system (1), with the modifications in blue,

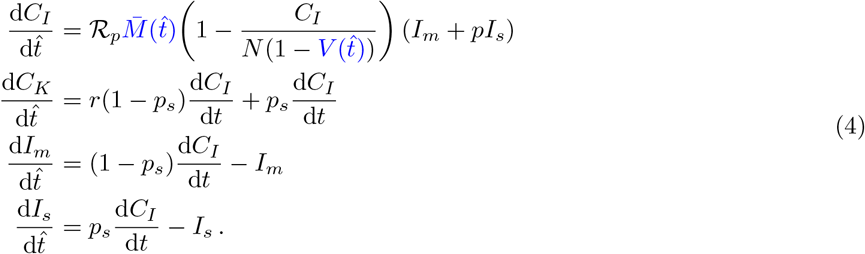

Here, *V*(*t*) is a non-negative monotonically increasing function of time bounded above by 1 which represents the total proportion of the population which is vaccinated. We fit the function

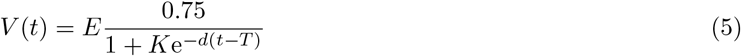

to vaccination administration data from [4]. We set the numerator to 0.75 as this is the estimated percentage of Canadians who are willing and able to get a vaccine [23]. *E* is an efficacy parameter. We set this to a conservative 0.6 as most of the Canadian population received a first dose of vaccine by the end of May, which has limited efficacy. This gives us a realistic ‘worst-case scenario’ for the fourth wave in Fall 2021. The parameters *K, d* and *T* are fit using vaccine administration data [4].

The new function 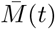 allows for explicit relaxation of non-pharmaceutical interventions beginning on day *τ* by using the functional form

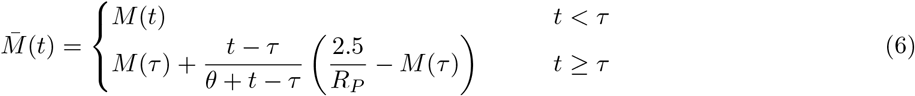

In the current study we consider the four largest provinces in Canada, British Columbia, Alberta, Ontario and Quebec. Fitted parameters for Model (1) and Equation 5 for each province are listed in Table 1.

**Table 1:** Table of fitted parameters for four provinces in study. Thee parameters *K, d* and *T* are the product of a single least-squares fit and thus do not have error bounds. Note that ℛ_*p*_ is the population reproduction number as of the start date of the fitting February 15, 2021 for Ontario and Quebec, and March 1, 2021 for Alberta and British Columbia.

| Province | Parameter | Mean ± Std. Dev. | Province | Parameter | Mean ± Std. Dev. |
| --- | --- | --- | --- | --- | --- |
| **British Columbia** | $\mathcal{R}_p$ | $1.93 \pm 0.86$ | **Alberta** | $\mathcal{R}_p$ | $0.44 \pm 0.72$ |
| | $p$ | $0.26 \pm 0.35$ | | $p$ | $0.33 \pm 0.39$ |
| | $r$ | $0.20 \pm 0.32$ | | $r$ | $0.24 \pm 0.33$ |
| | $k$ | $0.70 \pm 0.25$ | | $k$ | $0.53 \pm 0.32$ |
| | $m$ | $5.44 \pm 4.32$ | | $m$ | $6.84 \pm 4.10$ |
| | $A$ | $3.8 \pm 3.67$ | | $A$ | $3.40 \pm 3.33$ |
| | $m_1$ | $704.02 \pm 26079$ | | $m_1$ | $0.86 \pm 0.33$ |
| | $n$ | $8.04 \pm 8.94$ | | $n$ | $4.59 \pm 1.72$ |
| | $K$ | 0.03411 | | $K$ | 0.0481 |
| | $d$ | 0.436 | | $d$ | 0.4469 |
| | $T$ | 15.39 | | $T$ | 14.35 |
| **Ontario** | $\mathcal{R}_p$ | $1.38 \pm 0.73$ | **Quebec** | $\mathcal{R}_p$ | $0.89 \pm 0.85$ |
| | $p$ | $0.017 \pm 0.082$ | | $p$ | $0.098 \pm 0.22$ |
| | $r$ | $0.010 \pm 0.098$ | | $r$ | $0.048 \pm 0.20$ |
| | $k$ | $0.50 \pm 0.27$ | | $k$ | $0.59 \pm 0.30$ |
| | $m$ | $6.94 \pm 4.02$ | | $m$ | $7.60 \pm 3.57$ |
| | $A$ | $(0.591 \pm 9.65 \times 10^{-5})$ | | $A$ | $(0.103 \pm 1.21 \times 10^{-7})$ |
| | $m_1$ | $7.09 \pm 1.41$ | | $m_1$ | $7.83 \pm 2.20$ |
| | $n$ | $35.8 \pm 6.46$ | | $n$ | $38.9 \pm 7.28$ |
| | $K$ | 0.046 | | $K$ | 0.0404 |
| | $d$ | 0.435 | | $d$ | 0.4417 |
| | $T$ | 14.71 | | $T$ | 14.68 |

### 2.2 Clinical Pathways Model

The clinical pathways model is informed by an underlying epidemiological model which must have cases separated by into mild and severe infections; these are defined as cases with no/mild/moderate symptoms, and severe symptoms needing hospitalization, respectively.

The clinical pathways module determines the demand for healthcare consultations (General Practitioners (GP), emergency departments (ED), and assessment clinics) and hospital beds (ward and ICU) given the daily case incidence provided by the epidemiological model. The modules were first developed by [33] – a detailed description of the module is provided in [33], with links to a github repository. A flow diagram is shown in Figure 2. Daily presentations (*α*) are divided into mild and severe classes, where it is assumed that *η* = 0.1 are severe [33, 12]. Severe cases are then evenly distributed into two presentation classes, early and late, that determine when they present to healthcare. It is assumed that 80% and 20% of mild cases will report to GPs and EDs, respectively, and that all severe cases will report to GPs or EDs, with proportions 80% and 20% if they report early, and 0% and 100% if they report late [33]. Additionally, if assessment clinics are provided, it is assumed that some mild and severe cases will present to these locations (25% of mild cases, 50% of severe cases [33]). The number of consultations in each setting is limited (see Table 2 and more detail below). It is assumed that, if a consultation is requested within one healthcare setting, but not available, the individual will try for a consultation in a different healthcare setting given by the following flow: assessment clinics *→*ED *→*GP. Futhermore, it is assumed that if a GP consult cannot be obtained, the individual will return home and will be successful in obtaining a telehealth consult on that particular day. Additionally, it is assumed that for mild cases, 10% of GP consults and 5% of ED consults will try to revisit EDs and GPs, respectively, the next day [33].

**Table 2:**
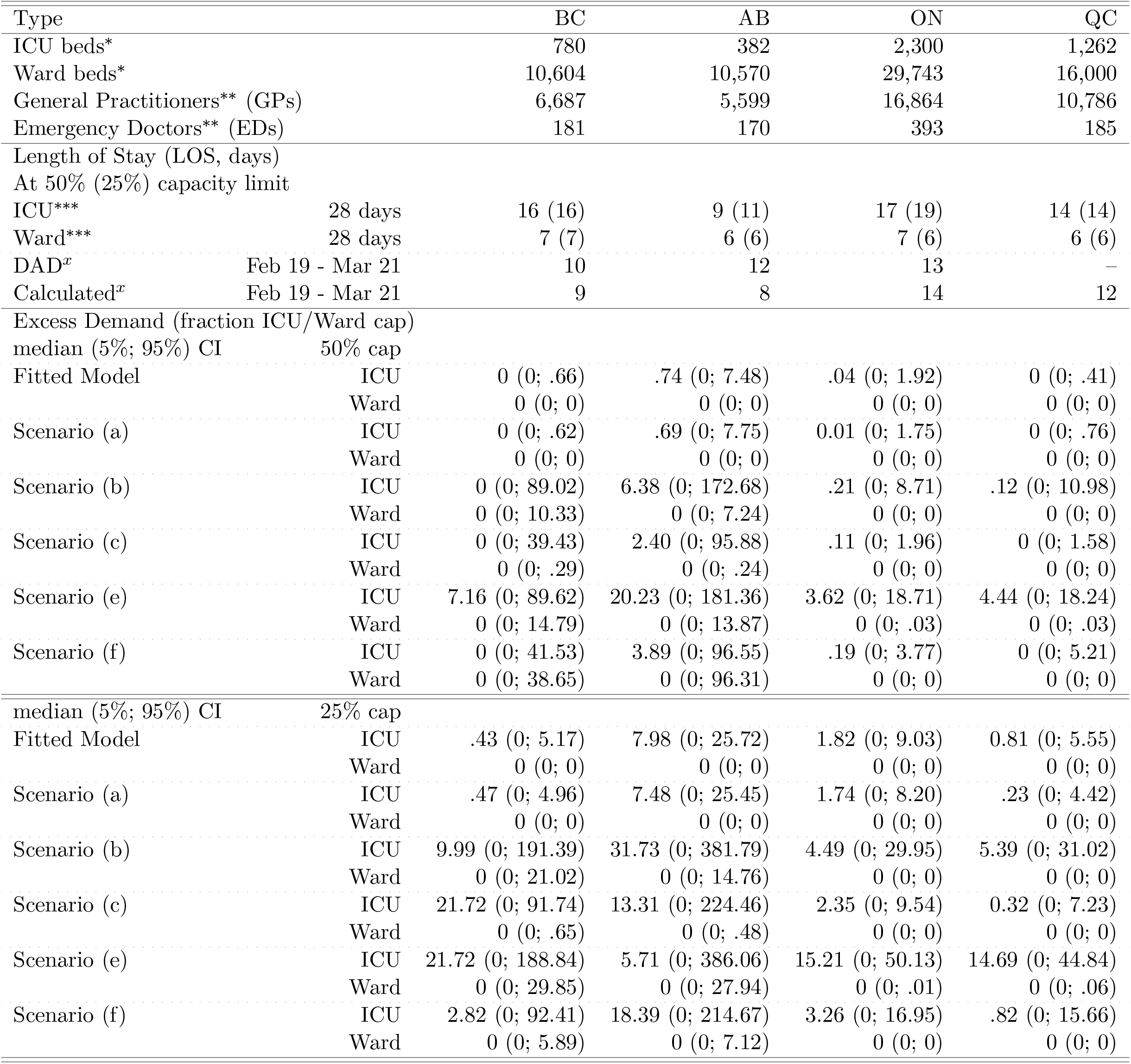
Healthcare bed and consult capacities, and length of stay, by province. Estimated from *[25, 17, 9] and **[13, 10]. *** calculated using provincial hospital data [11]. ^*x*^calculated from [**?**] and from model simulation output, from Feb 19 to Mar 21 2021. Excess demand for ICU and Ward beds for the fitted model, and Scenarios (a-c), (e-f).

**Figure 2:**
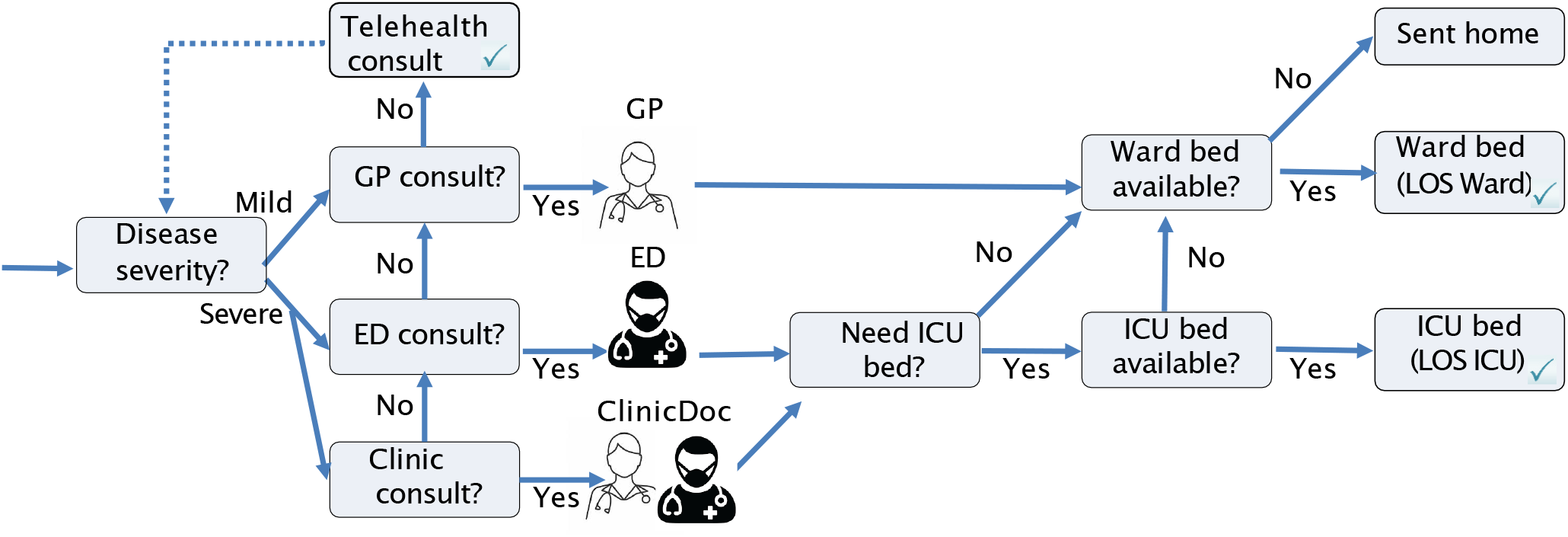
Clinical Pathways Flow Diagram. Adapted from [33].

Severe cases are considered for hospital admission. If a hospital bed is available, a severe case will be admitted. Probabilities of hospitalization given infection, by age, can be used, though we ignore age distribution in the current study given that our epidemiological model is not age stratified. Hospital beds are allocated depending on the location of consultation - first assessment clinics, then EDs, then GPs. Upon admission, the type of hospital bed is determined. Approximately 30% of all admissions will request an ICU bed [12]. If an ICU bed is needed but is not available, a ward bed will be allocated. However, if a ward bed is needed and is not available, the individual will be sent home.

GP, ED, and assessment clinic consultation availabilities, and hospital bed capacities are informed by national and provincial reports [10, 13, 12]. See Table 2. Similar to Moss et al. [33], we assume that 50% or 25% of the total capacity of GP and ED consults, and ICU and ward beds can be devoted to COVID-19 by default. It is assumed that assessment clinics are fully devoted to COVID-19, but that the provision of assessment clinics reduces the number of consultations available by GPs and EDs, as GP and ED doctors are needed to work within the assessment clinic setting. Similar to Moss et al. [33], we assume that 10% of the GP and ED workforce are needed to staff assessment clinics.

Once admitted to hospital, it is assumed that a patient will stay in the hospital for a particular length of stay (LOS). The LOS can depend on the province of study, but it can also change over time given changes in disease severity from dominant viral strains. In the current study, we choose to consider LOS’s of 4-19 days in ward beds, and 8-40 days for ICUs [12]. We however assume that the ward bed LOS must be less than that of the ICU LOS.

## 3 Results

Figures 3, 4, 5, 6 show the fit of Model (4) to cumulative and daily incidence data for British Columbia, Alberta, Ontario and Quebec, respectively (all subplots, blue line). In each figure, we also plot six different scenarios, considering relaxation in public health interventions and modifications to vaccine efficacy. For these scenarios, we switch from Model (1) to (4), with Equations (5) and (6) in early June 2021. In Scenario (a), we assume that NPI relaxation does not occur (*τ* =∞), and we assume that *V*(*t*) = 0. Here, the projection using Model (4) (pink line) agrees with the fitted Model (1) (blue line). In Scenario (b), we allow for NPI relaxation, starting June 1, 2021 and assuming that *θ* = 10, but we again assume that *V*(*t*) == 0. This allows us to quantify the effects of relaxation in the system. In Scenarios (b) and (c), we implement both equations (5) and (6) (with *θ* = 10), again with NPI relaxation starting on June 1. We, however, assume that the vaccine efficacy is higher in Scenario (d), changing *E* = 0.6 to *E* = 0.8. Comparing Scenarios (b), (c), and (d), we observe that vaccination reduced the impact of NPI relaxation, and this is more pronounced as vaccine efficacy increases (infection incidence in subplot (d) is less than that in subplot (c)). Finally, in Scenarios (e) and (f), we consider the same conditions as in Scenarios (b) and (c), but we increase the effect of NPI relaxation, decreasing *θ* from 10 to 1. Intuitively, we find a larger resurgence in Fall 2021 in Scenarios (e) and (f).

**Figure 3:**
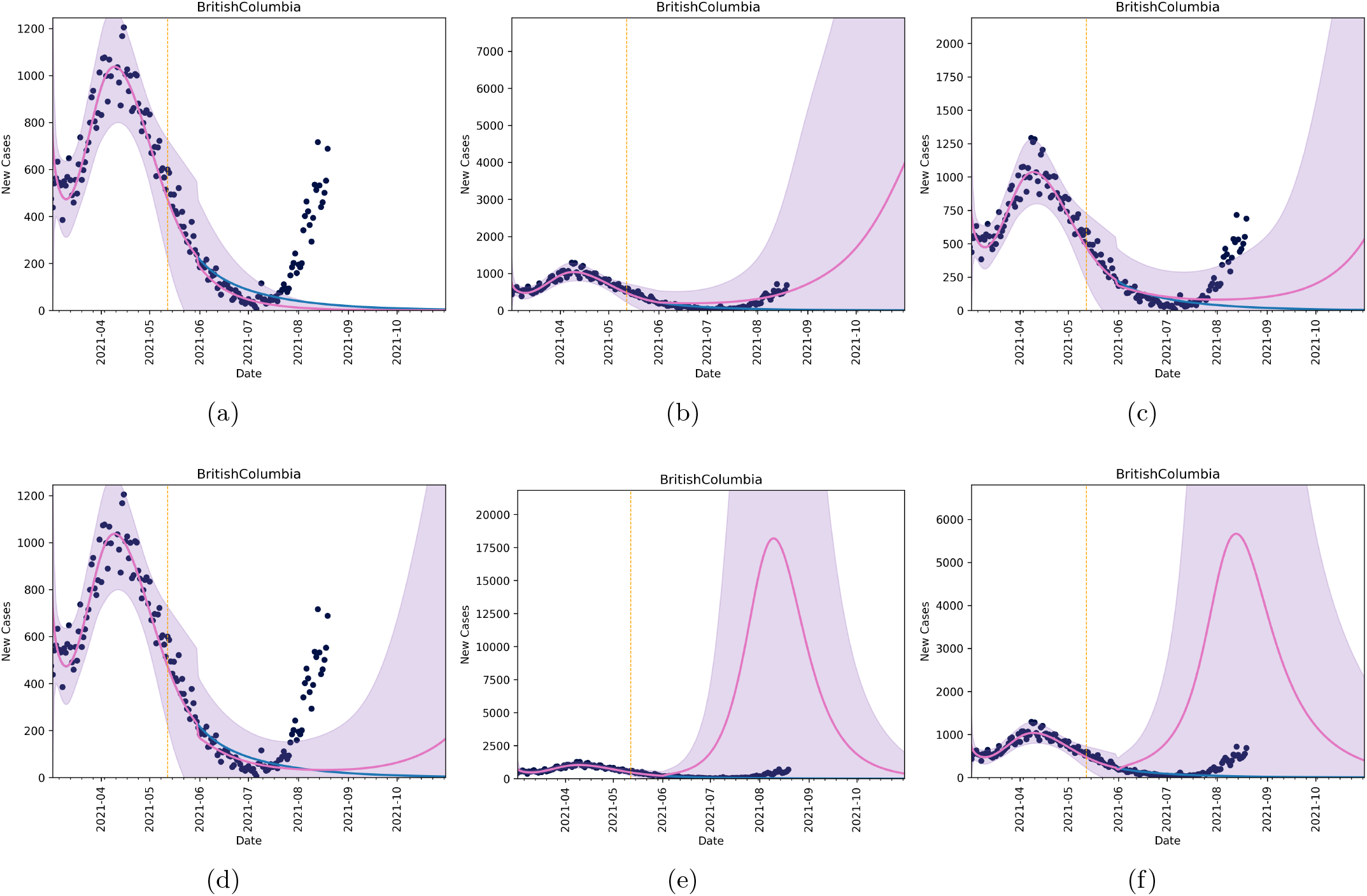
Forecasts in British Columbia under six different scenarios. In all scenarios, the blue line corresponds to Model (1) with equation (2). Black dots are data and the vertical line is the last date used for model fitting. The pink line (and purple shaded region) correspond to the mean and confidence intervals for six different scenarios under Model (4), with equations (5) and (6). The scenarios are implemented in early June. (scenario a) no NPI relaxation, *τ* = ∞, (scenario b) no change in vaccination, *V*(*t*) = 0, and NPI relaxation with *θ* = 10, (scenario c) a combination of vaccination and relaxation assuming starting in early June, (scenario d) the same as in scenario (c), but with *E* = 0.8. Scenario (e) shows relaxation with no vaccination with parameter *θ* = 1. Scenario (f) shows the combined effects of vaccination and relaxation with *E* = 0.6 and *θ* = 1.

**Figure 4:**
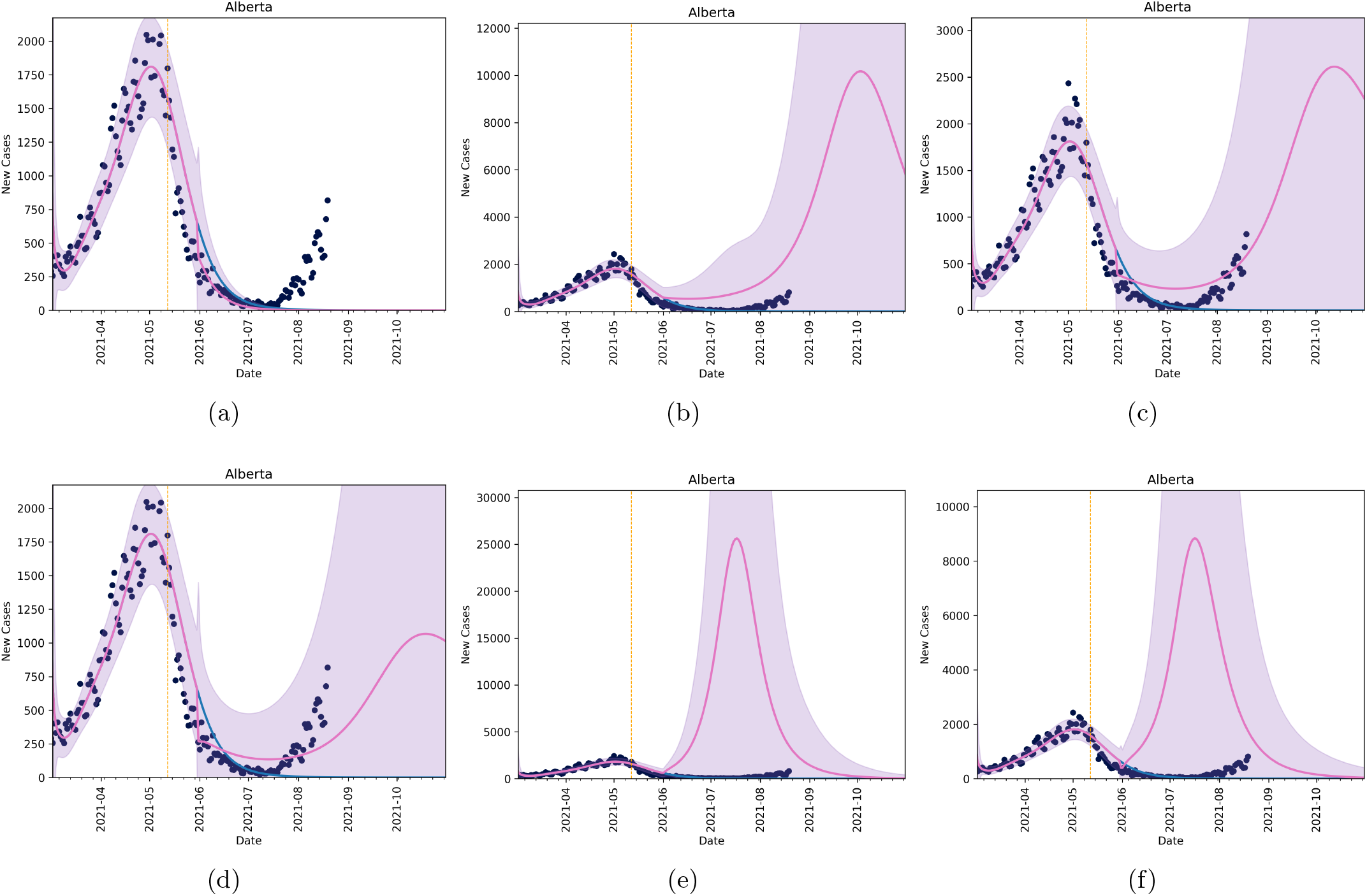
Forecasts in Alberta under four different scenarios. In all scenarios, the blue line corresponds to Model (1) with equation (2). Black dots are data and the vertical line is the last date used for model fitting. The pink line (and purple shaded region) correspond to the mean and confidence intervals for six different scenarios under Model (4), with equations (5) and (6). The scenarios are implemented in early June. (scenario a) no NPI relaxation, *τ* = ∞, (scenario b) no change in vaccination, *V*(*t*) = 0, and NPI relaxation with *θ* = 10, (scenario c) a combination of vaccination and relaxation assuming starting in early June, (scenario d) the same as in scenario (c), but with *E* = 0.8. Scenario (e) shows relaxation with no vaccination with parameter *θ* = 1. Scenario (f) shows the combined effects of vaccination and relaxation with *E* = 0.6 and *θ* = 1.

**Figure 5:**
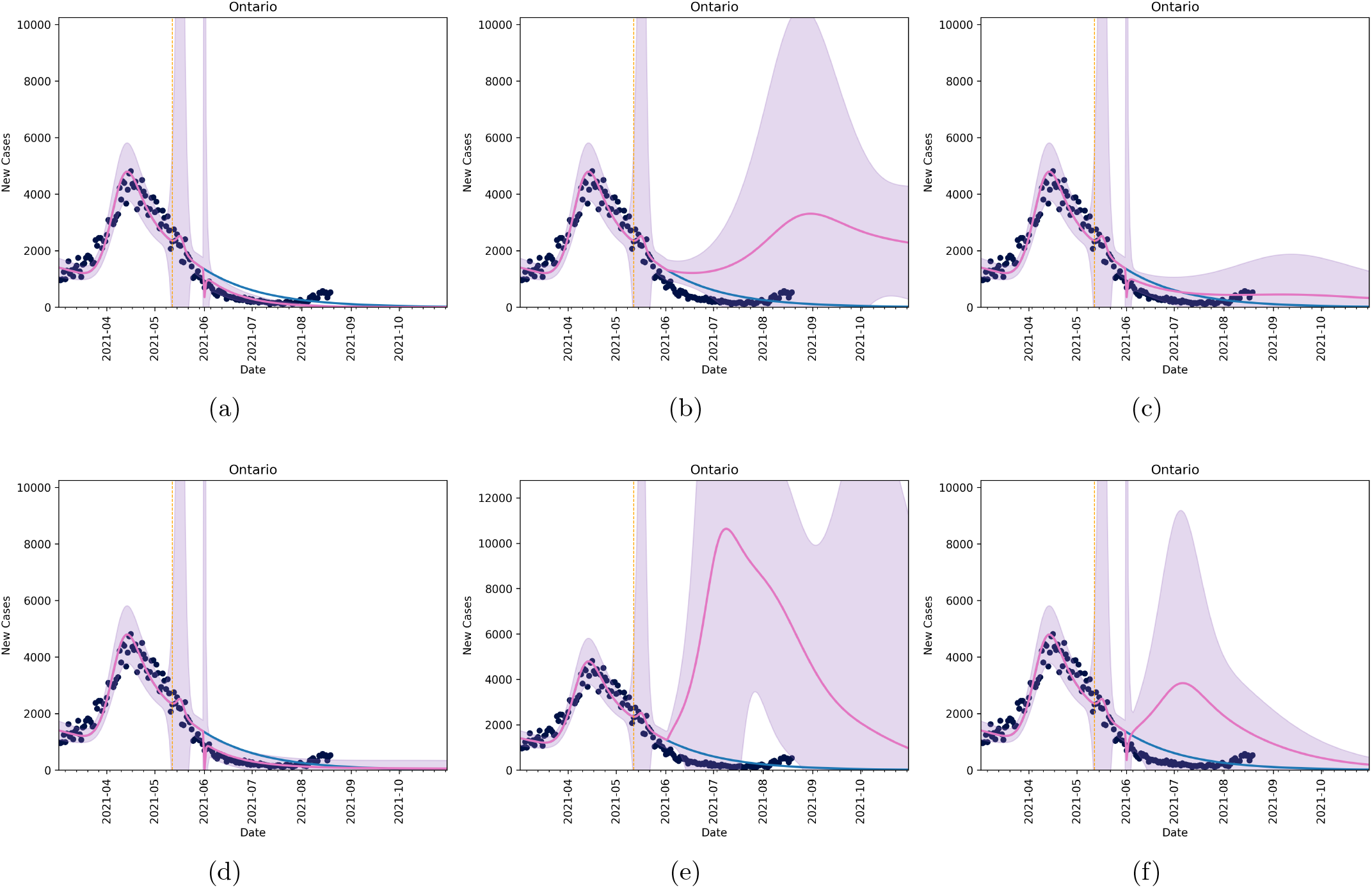
Forecasts in Ontario under six different scenarios. In all scenarios, the blue line corresponds to Model (1) with equation (2). Black dots are data and the vertical line is the last date used for model fitting. The pink line (and purple shaded region) correspond to the mean and confidence intervals for six different scenarios under Model (4), with equations (5) and (6). The scenarios are implemented in early June. (scenario a) no NPI relaxation, *τ* = ∞, (scenario b) no change in vaccination, *V*(*t*) = 0, and NPI relaxation with *θ* = 10, (scenario c) a combination of vaccination and relaxation assuming starting in early June, (scenario d) the same as in scenario (c), but with *E* = 0.8. Scenario (e) shows relaxation with no vaccination with parameter *θ* = 1. Scenario (f) shows the combined effects of vaccination and relaxation with *E* = 0.6 and *θ* = 1.

**Figure 6:**
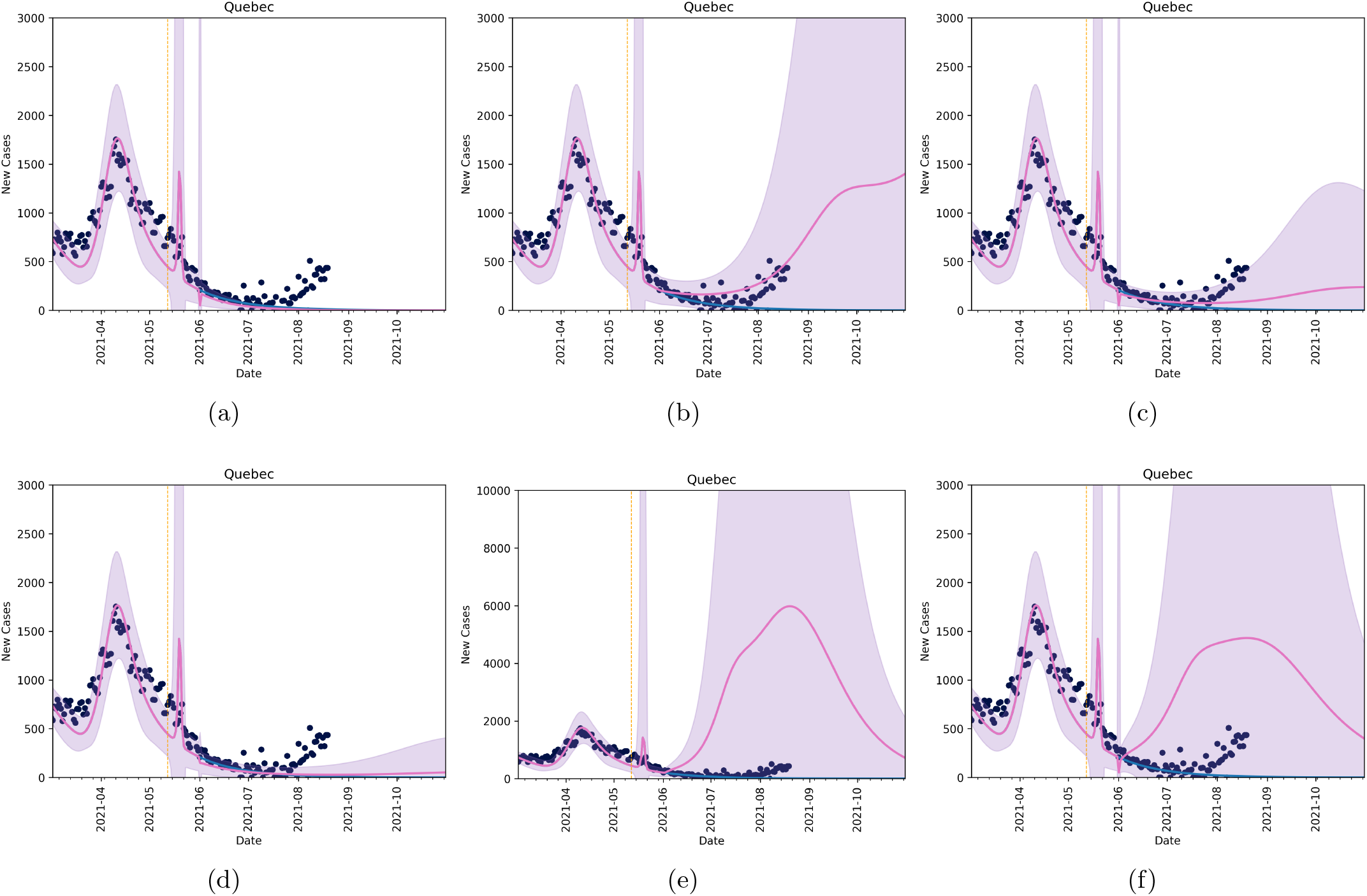
Forecasts in Quebec under six different scenarios. In all scenarios, the blue line corresponds to Model (1) with equation (2). Black dots are data and the vertical line is the last date used for model fitting. The pink line (and purple shaded region) correspond to the mean and confidence intervals for six different scenarios under Model (4), with equations (5) and (6). The scenarios are implemented in early June. (scenario a) no NPI relaxation, *τ* = ∞, (scenario b) no change in vaccination, *V*(*t*) = 0, and NPI relaxation with *θ* = 10, (scenario c) a combination of vaccination and relaxation assuming starting in early June, (scenario d) the same as in scenario (c), but with *E* = 0.8. Scenario (e) shows relaxation with no vaccination with parameter *θ* = 1. Scenario (f) shows the combined effects of vaccination and relaxation with *E* = 0.6 and *θ* = 1.

For comparison, we plot the daily incidence data from June 1 to Aug 15 2021 against all scenarios (a) to (f). It is clear from these figures that scenario (b) or (c) produce mean daily incidence numbers of similar magnitude to the most recent case report data. Considering a best scenario, scenario (c), produces incidence numbers that capture incidence numbers for all provinces in the 95 confidence interval (pink shaded area).

### 3.1 Length of Stay

The model fits for each province of study were introduced into the healthcare demand module to determine ICU and Ward bed lengths of stay (LOS) that best match provincial COVID-19 hospital bed occupancy data [11] for the entire month of May 2021. Here, we considered availability matching 50% and 25% capacity of the ICUs and Wards in each province. The resulting ICU and Ward bed LOS’s under both assumptions are listed in Table 2, Fitted Model.

The Canadian Discharge Abstract Database (DAD) records the LOS for every patient entering and exiting all ICU beds in every province and territory (Quebec is not required to report). Table 2 lists the average ICU LOS calculated from Feb 19 to Mar 21 2021. We have also included and calculated ICU LOS from our model for the same time period. They are in close agreement.

### 3.2 Excess Demand

High case numbers can result in excess demand for ICU and Ward beds. The healthcare demand model calculates the number of ICU and Ward bed admissions that have not been met for each relaxation scenario considered above (except Scenario (d)). Table 2 provides the median and (5%, 95%) confidence intervals for the excess demand for each province given the corresponding ICU and Ward bed LOS’s, assuming 50% and 25% availability for COVID patients. Considering a maximum capacity of 50% of the total beds, we observe that the median ICU excess demand reaches a number greater than zero for BC under Scenario (e) only. All scenarios have excess ICU demand for the provinces of Alberta and Ontario, and Quebec incurs excess ICU demand under scenarios (c) and (f) only. We however also observe that the median Ward bed excess demand for all scenarios, for all provinces, is zero for all scenarios. This means that there is sufficient Ward bed availability to accommodate all patients turned away from the ICU, and accommodate all admissions to Ward beds directly from the GP, Clinics, and ED consultations for each of the provinces. Therefore, there is no excess demand for Ward beds and the Fall 2021 resurgence predicted by the model under a 50% cap of ICU and Ward beds. Considering a cap of 25%, intuitively, the excess demand increases. Our models results again show that median excess ward demand is zero.

Figures 7 and 8 plot the daily admissions (left column) and occupancy (right column) of the ICU (top column) and ward (bottom row) beds for each province for the respective LOS’s, assuming 50% and 25% maximum bed availability, considering Scenario (e) conditions (a scenario with high excess demand for all provinces under study). Here, we observe that the Fall 2021 wave will reach high enough levels such that ICUs in each province will reach capacity - a plateau in admission and occupancy levels occurs. We, however, also observe that, on average (mean = red line, median = black line), the Ward bed capacity is not achieved in any of the four provinces.

**Figure 7:**
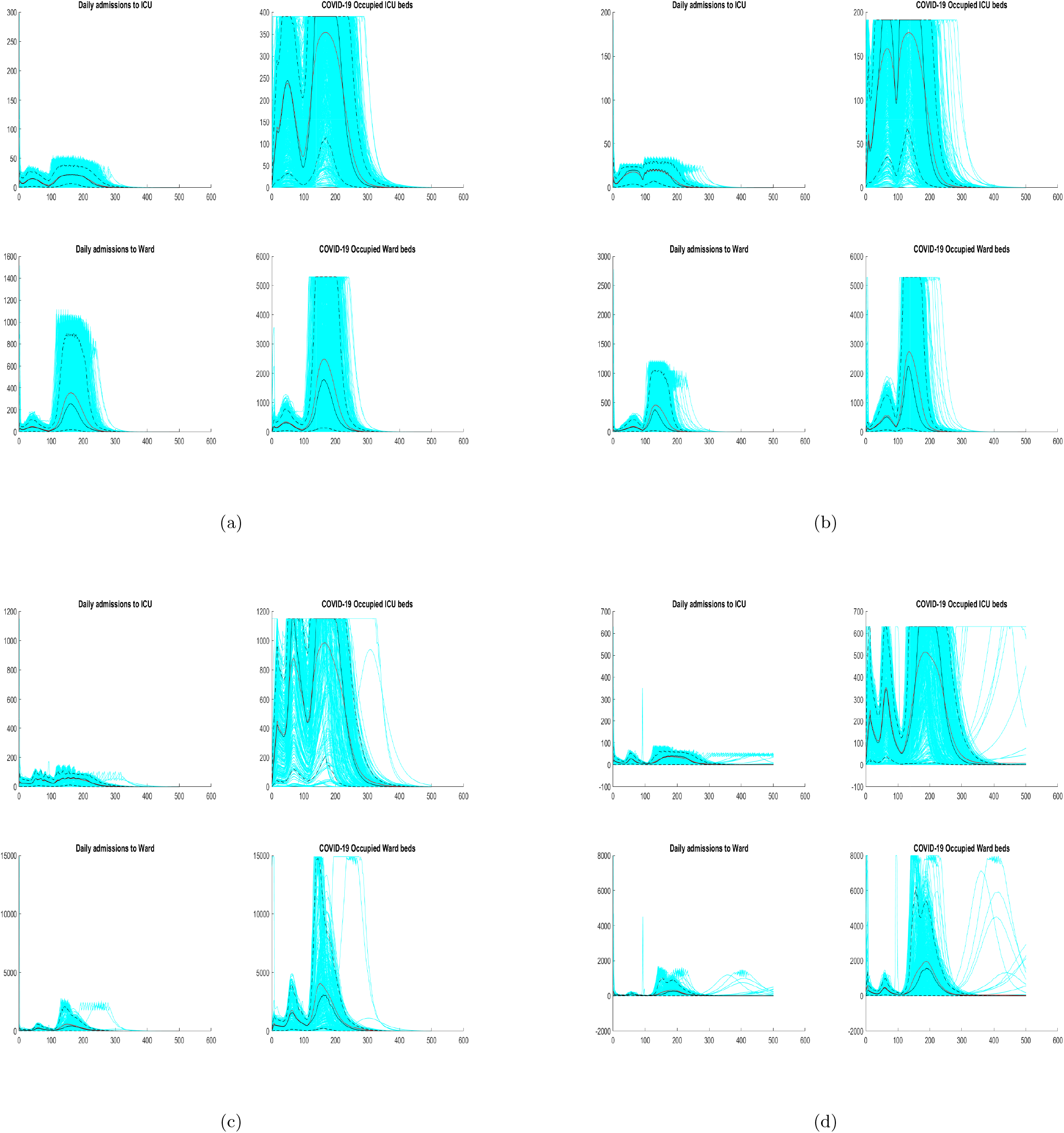
Daily admission and occupancy of ICU and Ward beds, by province, assuming 50% maximum bed availability, for Model (4), with Equations (5) and (6), with *E* = 0.6 and *θ* = 1 − Scenario (e). (panel a) BC, (panel b) AB, (panel c) ON, (panel d) QC. The daily admissions (left column) and occupancy (right column) of ICU (top row) and Ward (bottom row) beds is shown for the provincial best match LOS - see Table 2. Results from April 1 to December 31, 2021 are shown. Admissions for each simulation are shown (light blue lines), with median (black line) and mean (red line).

**Figure 8:**
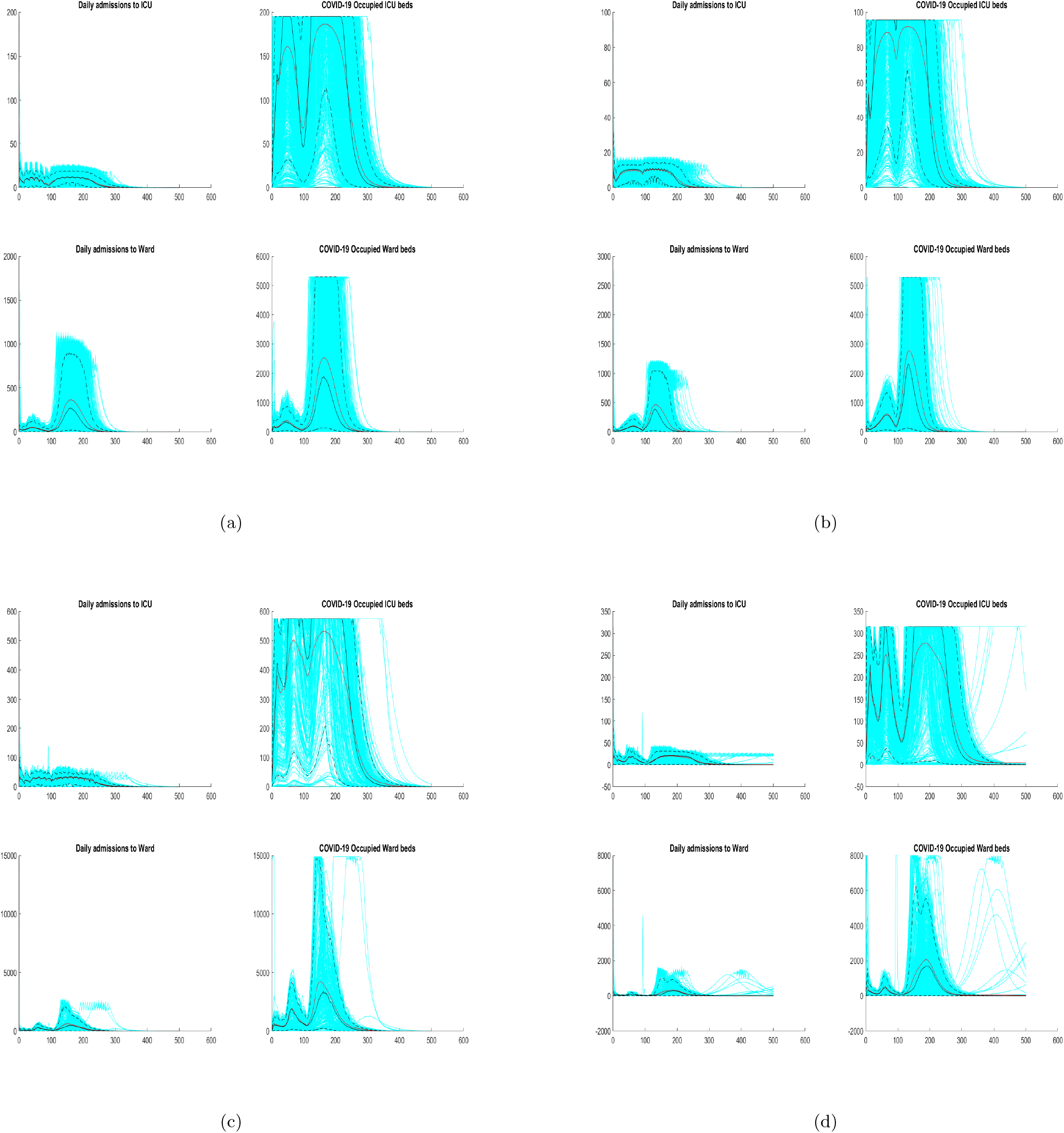
Daily admissions and occupancy of ICU and Ward beds, by province, assuming 25% maximum bed availability, for Model (4), with Equations (5) and (6), with *E* = 0.6 and *θ* = 1 − Scenario (e). (panel a) BC, (panel b) AB, (panel c) ON, (panel d) QC. The daily admissions (left column) and occupancy (right column) of ICU (top row) and Ward (bottom row) beds is shown for the provincial best match LOS - see Table 2. Results from April 1 to December 31, 2021 are shown. Admissions for each simulation are shown (light blue lines), with median (black line) and mean (red line).

## 4 Discussion

With the introduction of variant strains [21, 29] of COVID-19, coupled with vaccine hesitancy [19] it is important to look to the future with a cautious lens. Here, we study the effects of an outbreak in different jurisdictions in Canada on the healthcare system. We estimate current length of stay in hospital for COVID-19 patients, and project forward into Fall 2021. A key result is that some jurisdictions in Canada may be at risk of straining their healthcare system in the fall as vaccine efficacy against variant strains of SARS-CoV-2 falls and vaccine uptake saturates.

Of course, the easiest ways to reduce this burden are to lower the average length of stay, which is not feasible unless there are new therapeutics, to increase the number of hospital ICU and Ward beds, something that can be managed but only to a limited extent, or to reintroduce non-pharmaceutical interventions. Non-pharmaceutical interventions such as lockdown will be increasingly hard to implement as frustration and fatigue of the population continue to rise [18]. Luckily, our results show that many of the most populous provinces will show little to no overburdening of the healthcare system in many of our scenarios.

While vaccination deployment has been successful and rapid in 2021 in Canada, there is still a careful balance that needs to be struck when considering relaxation of public health measures. Relaxation too quickly can quickly create a situation where vaccination efforts are undermined [6, 5] and the consequences fall on the healthcare system. The model predicts that various provincial healthcare services may not see overall excess demand in Fall 2021, but could still see an overburden on ICU needs. While the model-predicted future of COVID-19 in Canada thus seems optimistic, it must be noted that Fall resurgence outcomes that do not see excess ICU demand should still be a goal for all Canadians. Increased vaccine uptake should thus be considered, as well as, increased uptake and proper practice of personal protective behaviours.

In this study we have employed a mathematical modelling framework to project COVID-19 epidemic scenarios and quantify healthcare demand. This framework can be modified to consider other infectious diseases. It can also be refined to consider larger and smaller jurisdictions. Refinement to particular Canadian healthcare regions is a course for future work.

## Data Availability

Data is available publicly or by request.

## Notes

### Competing Interest Statement

The authors have declared no competing interest.

### Funding Statement

Funded by NSERC discovery grants of Matthew Betti and Jane Heffernan.

